# NHS CHECK: protocol for a cohort study investigating the psychosocial impact of the COVID-19 pandemic on healthcare workers

**DOI:** 10.1101/2021.06.08.21258551

**Authors:** Danielle Lamb, Neil Greenberg, Matthew Hotopf, Rosalind Raine, Reza Razavi, Rupa Bhundia, Hannah Scott, Ewan Carr, Rafael Gafoor, Ioannis Bakolis, Siobhan Hegarty, Emilia Souliou, Anne Marie Rafferty, Rebecca Rhead, Danny Weston, Sam Gnangapragasam, Sally Marlow, Simon Wessely, Sharon A.M. Stevelink

## Abstract

**Introduction:** The COVID-19 pandemic has had profound effects on the working lives of healthcare workers (HCWs), but the extent to which their well-being and mental health have been affected remains unclear. This longitudinal cohort study aims to recruit a cohort of NHS healthcare workers, conducting surveys at regular intervals to provide evidence about the prevalence of symptoms of mental disorders, investigate associated factors such as occupational contexts and support interventions available.

**Methods and Analysis:** All staff, students, and volunteers working in each of the 18 participating NHS Trusts in England will be sent emails inviting them to complete a survey at baseline, with email invitations for the follow up surveys being sent 6 and 12 months later. Opening in late April 2020, the baseline survey collects data on demographics, occupational and organisational factors, experiences of COVID-19, a number of validated measures of symptoms of poor mental health (e.g. depression, anxiety, post-traumatic stress disorder; PTSD), and measures of constructs such as resilience and moral injury. These regular surveys will be complemented by in-depth psychiatric interviews with a select sample of healthcare workers. Qualitative interviews will also be conducted, to gain deeper understanding of the support programmes used or desired by staff, and facilitators and barriers to accessing such programmes.

**Ethics and Dissemination:** Ethical approval for the study was granted by the Health Research Authority (reference: 20/HRA/210, IRAS: 282686) and local Trust Research and Development approval. Cohort data are being collected via Qualtrics online survey software, are pseudonymised and held on secure University servers. Participants are aware that they can withdraw from the study at any time, and there is signposting to support services for any participant who feels they need it. Only those consenting to be contacted about further research will be invited to participate in the psychiatric and qualitative interview components of the study. Findings will be rapidly shared with NHS Trusts to enable better support of staff during the pandemic, and via academic publications in due course.

**Strengths and limitations of this study:**

- The longitudinal cohort design addresses the lack of long-term data on this population, and the current predominance of cross-sectional evidence available.
- The availability of Trust HR data means we will be able to calculate response rates, and weight the data appropriately.
- The diagnostic interview component of the study will allow us to establish the true prevalence of mental disorders, which can be inflated by the measures used in most mental health and wellbeing cohort studies.
- The qualitative interviews will give deeper insight into the support programmes that HCWs find most helpful, and provide ideas for Trusts to improve their offer to staff.
- The three components of the study give breadth and depth lacking in much of the mental health and wellbeing research currently available, but there is a risk of over-burdening already stretched HCWs.

## Introduction

The COVID-19 pandemic raises many questions on biological, behavioural, emotional and social responses to a global threat.[1] Evidence from systematic reviews and meta-analyses suggests that healthcare workers who have had to deal with serious infection during pandemics in the past (SARS, MERS, Ebola, swine flu) are at increased risk of both current and subsequent mental health problems.[2,3]

Pandemics expose healthcare workers to overwork, isolation from friends and family, discrimination, exhaustion and an increased risk of developing common mental disorders.[3] Early accounts from Wuhan, China, confirm this, suggesting that the added pressures of providing healthcare during a pandemic can result in impaired decision making, attention and understanding thereby hindering the control of the pandemic, but also early signs of distress may well lead to longer term mental ill-health.[4] A relatively new concept that has attracted a lot of attention is the concept of ‘moral injury’.[5] Moral injury describes the psychological distress resulting from actions, or the lack of them, which violate someone’s moral or ethical code and can contribute to the development of mental health difficulties, including depression, post-traumatic stress disorder (PTSD) and suicidal ideation.[6].. During the COVID-19 pandemic many National Health Service (NHS) staff may have to make difficult choices not faced before, to deliver care that they know is suboptimal, and have to explain difficult decisions to relatives. Such ethical dilemmas will be new, in scale and nature.

Research to date suggests that women, younger people, and those from racial and ethnic minority groups are at higher risk of adverse outcomes.[7] There is evidence that those in lower income brackets have been more negatively affected,[8] and that nurses may be worse affected than those in other roles.[9] In terms of trajectory, wellbeing appears to have worsened during the early stages of the pandemic, with small improvements through the following months.[8] Other factors associated with poor mental health and wellbeing are lack of access to Personal Protective Equipment (PPE) REF, and lack of supportive environments.[10] While research on moral injury has previously focussed on military contexts, early work in healthcare settings shows concerning associations with poorer outcomes.[10,11]

Much of the existing research in this area is based on single questionnaire assessments, typically only including clinical staff rather than all healthcare workers, from which bold claims of mental health crisis are made. While survey data can be informative, two stage epidemiological studies using diagnostic interviews tend to show that such surveys over-estimate prevalence of mental disorders.[12] To address these issues, our study has five distinct features: (1) breadth geographically, with a range of different types of NHS Trust (e.g. acute, mental health) and locations (urban, rural, all areas of England); (2) depth phenotypically, with diagnostic interviews included in order to ascertain true prevalence of mental disorders rather than simply indicators of distress; (3) longitudinal data collection to look at temporal patterns, meaning we will be able to identify whether a surge of symptoms at time of crisis lead to persistence or remission; (4) inclusivity regardless of role, meaning that we will include all healthcare workers who are contributing to the pandemic effort, whether they are in clinical or ancillary and support roles; and (5) the ability to calculate accurate response rates and weight data appropriately, thanks to demographic population data provided by each participating Trust’s Human Resources (HR) department.

### Aims, objectives, and hypotheses

The main aim of the study is to investigate the psychosocial impacts of the COVID-19 pandemic on NHS Trust workforce mental health and wellbeing over time. In addition, we aim to explore the uptake and usefulness of staff support interventions available to participants.

The primary objective is to establish a cohort of all staff employed in participating NHS Trusts, to carry out repeated surveys of their mental health and wellbeing and psychiatric diagnostic interviews to determine the true prevalence of disorders. Based on the literature outlined above, we have a number of descriptive aims and hypotheses. We will:

1. Describe the prevalence of psychological distress and characteristics associated with poorer mental health. Hypotheses include:
  ⍰ *Poorer mental health will be associated with demographic variables (including younger age, female sex, coming from Black, Asian, or other racial or ethnic minority groups)*.
  ⍰ *Poorer mental health will be associated with occupational characteristics (including role, pay grade, work setting, redeployment status)*.
2. Establish the true prevalence of mental disorders via diagnostic interviews. Secondary objectives include exploring the factors associated with poor mental health, patterns of reported distress over time, and qualitatively exploring experiences of staff support interventions. We will:
3. Describe workplace factors associated with poorer mental health. Hypotheses include:
  ⍰ *Poorer mental health will be associated with higher levels of reported moral injury*.
  ⍰ *Poorer mental health will be associated with reported lack of access to personal protective equipment (PPE)*.
  ⍰ *Poorer mental health will be associated with perceived lack of support from leaders, team, friends/family*.
4. Describe patterns and persistence of psychological distress symptoms over time. Hypotheses include:
  ⍰ *Mental health and wellbeing will follow the trajectory of the pandemic, with poorer outcomes evident during/after higher levels of COVID-19 prevalence (e.g. as measured by daily deaths, hospital admissions)*.
  ⍰ *Poorer mental health at baseline will be associated with poorer mental health at 6 and 12 month follow up points*.
  ⍰ *Predictors of poorer mental health at follow up time points will include younger age, female sex, racial or ethnic minority group, being a nurse, perceived lack of support, lack of access to personal protective equipment (PPE), and higher levels of reported moral injury*.
5. Qualitatively evaluate tiered and tailored staff support programmes being implemented locally and nationally, using in-depth interviews and thematic analysis. This will enhance our understanding of how we could further scale up effective support programmes within and across Trusts.
6. Provide a platform for further randomised controlled trials (RCT), observational, or intervention studies, including:
  ⍰ An ethnic inequalities module, which will explore ethnic inequalities in mental health and occupational outcomes across NHS staff and the mechanisms that perpetuate these inequalities, using a mixture of quantitative and qualitative data (the Tackling Inequalities and Discrimination Experiences in health Services study: TIDES https://tidesstudy.com).
  ⍰ Estimation of parameters of interest which could be later incorporated when designing an RCT or a pragmatic trial, e.g. prevalence of psychosocial distress, intra-class correlation coefficient across NHS trust.
  ⍰ An RCT testing a wellbeing app for use by NHS staff.

## Methods

### Design and setting

This study consists of three main components:

i. A longitudinal cohort study, consisting of surveys administered at baseline, 6 months, and 12 months.
ii. A diagnostic interview study, using a clinical diagnostic measure to ascertain the true prevalence of mental disorders.
iii. A qualitative interview study, using semi-structured interviews to explore experiences of using (or reasons for not using) staff support programmes.

The study will be carried out in 18 NHS Trusts in locations across England.

### Study population and sample

#### Longitudinal cohort study

Participants across the study will be any NHS-affiliated staff including clinical staff, students and all support staff working within participating NHS Trusts: Avon and Wiltshire Mental Health NHS Foundation Trust (N=4,334); Cambridge University Hospitals NHS Foundation Trust (N=10,243); Cambridgeshire and Peterborough NHS Foundation Trust (N=4,235); Cornwall Partnership NHS Foundation Trust (N=3,977); Devon Partnership NHS Foundation Trust (N=3,280); East Suffolk and North Essex NHS Foundation Trust (N=10,219); Gloucestershire Hospitals NHS Foundation Trust (N=8,437); Guys and St Thomas’ NHS Foundation Trust (N=19,760); King’s College Hospital & PRUH (N=12,959); Lancashire and South Cumbria NHS Foundation Trust (N=6,984); Norfolk and Norwich University Hospitals (N=10,502); Nottinghamshire Healthcare NHS Foundation Trust (N=8,860); Royal Papworth Hospital (N=2,110); Sheffield Health and Social Care (N=2,610); South London and Maudsley NHS Foundation Trust (N=5,151); Tees Esk and Wear Valleys NHS Foundation Trust (N=7,315); University Hospitals of Derby and Burton (N=13,231); University Hospitals of Leicester NHS Foundation Trust (N=16,946); and/or any of the Nightingale Hospitals (London, Exeter, Leeds/Harrogate, Cardiff, and Manchester), dependent on whether these sites are active at the time of data collection.

#### Diagnostic interview study

Participants will be up to 350 HCWs who have completed the baseline NHS CHECK survey.

Up to 250 participants will be purposively sampled according to their responses to the General Health Questionnaire in the short survey module;[13] half will meet caseness and half will not meet caseness. These participants will be administered the Clinical Interview Schedule-Revised (CIS-R).[14] The number of CIS-R interviews (n=250) has been chosen balancing precision and cost. Based on a simulation study, we anticipate that 250 interviews will allow estimation of CIS-R prevalence of common mental disorders to within plus or minus 6%.

Up to 100 HCWs will be sampled according to their responses to the Post Traumatic Stress Disorder Checklist civilian version [15] in the long survey module; half will meet caseness and half will not meet caseness. These participants will be administered the Clinician Administered PTSD Scale for DSM-5 (CSPS-5).[16] This sample size was calculated as the necessary number to achieve a valid estimation of PTSD prevalence.

Sampling for the CIS-R and CAPS-5 groups will be stratified to ensure representation from each of 18 participating sites in the NHS CHECK study, as well as to ensure ethnic, sex, and age breakdown that resembles respondents of the baseline survey.

#### Qualitative interview study

Participants will be sampled from two groups: i) Up to 48 participants who complete the NHS CHECK 6 month follow up survey; and ii) up to 12 members of staff (from participating Trusts) who are involved in implementing staff support programmes, who do not need to have completed any NHS CHECK surveys. Group i) will be sampled according to sex, ethnicity, age (a cut-off of 50 years will be used to dichotomise participants into younger or older groups), and occupational role. Balancing these demographic factors, four discrete groups of HCWs will be sampled, with 12 in each group: a) those who used support programmes and found them helpful; b) those who used support programmes but did not find them helpful; c) those who heard about support programmes but did not use them; and d) those who had not heard about support programmes. We will endeavour to include a diverse range of participants in group ii), but this will be dependent on those involved in staff support in participating Trusts.

Participants in Group i) will be recruited first, and what they tell us will inform subsequent interviews with those in Group ii).

### Inclusion criteria

Inclusion criteria for each part of the study are as follows.

#### Longitudinal cohort study

Participants must:

1. Be an NHS-affiliated member of staff, working at, or with (e.g. volunteer or student), the participating NHS Trusts and/or Nightingale Hospitals during the COVID-19 pandemic;
2. Be aged 18 and over;
3. Be able to give informed consent to take part in research;
4. Be able to understand and communicate in English;
5. Have access to an email address to facilitate survey registration and receive follow up survey links.

#### Diagnostic interview study

Participants must meet the criteria for the cohort study, and must also:

1. Have completed the baseline NHS CHECK survey as an NHS member of staff based at a participating Trust; including the relevant PTSD survey measures if being administered the CAPS-5.
2. Have indicated in the baseline survey that they consent to be contacted for participation in further research.
3. Have access to a phone for the interview.
4. Have scored ≥ 4 on the GHQ to meet caseness for probable common mental disorders or ≥ 14 on the PCL-6 to meet caseness for probable PTSD.

#### Qualitative interview study

Participants in group i) must meet the criteria for the cohort study, and must also:

1. Have completed the baseline and 6 month follow up surveys of the longitudinal cohort study;
2. Have indicated in the baseline survey that they consent to be contacted for participation in further research.
3. Have provided data on whether or not that have heard of and/or used any staff support programmes.

Participants in group ii) must:

1. Have been involved in implementing staff support programmes in one of the 18 participating Trusts or Nightingale hospitals.
2. Be aged 18 and over;
3. Be able to give informed consent to take part in research;
4. Be able to understand and communicate in English.

### Measures

#### Longitudinal cohort study

##### Baseline

The baseline survey will involve a short survey (5-10 minutes), which collects information on the following topics: 1) contact details, 2) occupational information (e.g. occupational group, length of professional registration), 3) socio-demographic characteristics, 4) working practices (e.g. access to personal protective equipment, performing aerosolising procedures), 5) perceived support from managers, colleagues, friends and family 6) COVID-19 related questions (e.g. suspected infection status, COVID-19 test status, isolation/quarantining), 8) staff support programme access, 9) self-reported diagnosed health conditions and 10) common mental disorders (CMDs). The prevalence of probable CMDs will be assessed using the 12-item General Health Questionnaire (GHQ-12), with a cut-off score of 4 or more indicating caseness.[13]

There will be the option for participants to complete an additional longer survey (10-15 minutes), which includes information on the following: 1) impact of COVID-19 (e.g. on family, income, health, positive and negative changes in personal life or work), 2) work experiences (leadership and teamwork, sickness absence, unsafe clinical practices, preparedness), 3) usefulness of staff support programmes, 4) caring responsibilities outside of work, 5) confidence in institutions to handle the COVID-19 pandemic. In addition, the following validated measures will be used in the longer survey: the 7-item Generalised Anxiety Disorder (GAD-7) scale to measure probable moderate anxiety disorder with a cut-off score of 10 or more indicating caseness;[17] the 9-item Patient Health Questionnaire (PHQ-9) to measure probable moderate depression with a cut-off score of 10 or more indicating caseness;[18] the 10-item Alcohol Use Disorder Identification Test to measure alcohol consumption with a cut-off score of 8 or more indicating hazardous drinking;[19] the 6-item Post-Traumatic Stress Disorder checklist (PCL-6) civilian version to measure post-traumatic stress disorder (PTSD) with a cut-off score of 14 or more indicating the presence of probable PTSD;[15] the 9-item Moral Injury Event Scale (MIES) to measure moral injury, with a higher score indicating greater exposure to morally injurious events;[20] the 14-item Warwick-Edinburgh Mental Well-being Scale (WEMWBS) to measure subjective well-being and psychological functioning, with a higher score indicating a higher level of mental well-being;[21] the 6-item Brief Resilience Scale (BRS) to measure psychological resilience, with a higher score indicating a higher level of resilience;[22] the 12-item Burn-out Assessment Tool (BAT) to measure burn out, with a cut-off score of 3.02 indicating probable burnout.[23]

We will also assess suicidal ideation in the longer survey, using items related to suicidal thoughts, suicide attempts and self-harm derived from the Clinical Interview Schedule (CIS-R).[14] We will measure fatigue using three novel items exploring: 1) emotional and physical exhaustion on a 0-10 scale; 2) whether any experienced fatigue is greater than usual tiredness (yes/no); and 3) whether any experienced fatigue interferes with ability to do things (yes/no).

##### Follow up – 6 months

The 6 month survey will also involve a short and long version. The short survey will collect information on the national and local staff support programmes used, and on household income. The same questions as at baseline will be asked regarding: COVID-19 experiences (e.g. suspected infection status, COVID-19 test status, isolation/quarantining); teamwork; support from colleagues and friends/family. The same measures will be used in the 6 month survey as at baseline, as follows: GHQ-12; GAD-7; PHQ-9; AUDIT; PCL-6; MIES; and the CIS-R suicidality questions. In addition, the short form of the Post-traumatic Growth Inventory (PTGI-SF will be used.[24]

The long survey will collect the same measures as at baseline: WEMWBS; BRS; BAT; Fatigue. It will also collect information on the personal and occupational factors as at baseline. The topics covered may be updated as the pandemic evolves, in order to capture the most relevant information.

##### Follow up – 12 months

Similar short and long versions will be used in the 12 month follow up survey as the 6 month survey, with further refinement as the pandemic evolves.

#### Diagnostic interview study

The Clinical Interview Schedule (Revised) (CIS-R) will be used to assess mental disorders.[14] The CIS-R is a standardised interview for use in general practice, community settings, hospitals, and occupational contexts, and consists of 10 items, each scored on a scale of 0-4, with scores combined to provide a total weighted score. Questions refer to symptoms in the previous week. Use of the CIS-R allows comparison with a national probability sampled survey, the Adult Psychiatric Morbidity Survey (APMS).[25]

The Clinician Administered PTSD Scale (CAPS-5) will be used to assess PTSD.[16] The CAPS-5 is a structured interview for use in clinical and research settings that assesses symptom severity and diagnostic status of PTSD in line with DSM-5 criteria. The tool consists of 30 items, on a 5 point rating system, across 7 criteria that ask about symptoms within the past month; scores are combined to create a total symptom severity score and used to establish presence or absence of the diagnosis.

#### Qualitative interview study

Semi-structured interview schedules will be constructed, using Normalisation Process Theory as a framework in order to address topics relevant to the evaluation and implementation of interventions.[26] Questions will be included to draw out details about: how and why staff support programmes have been used, including ease of access and use; whether they have been helpful or unhelpful, and why; how information about such interventions could be communicated to staff most helpfully; and whether there are additional supports that would be helpful for Trusts to provide.

### Study procedures

#### Longitudinal cohort study

##### Recruitment

Potential participants will be identified via each Trust Human Resources (HR) system. Senior management at each site will use existing dedicated group email lists to circulate details of the study and the URL for the baseline survey of the longitudinal cohort study, emphasising the voluntary nature of participation. A ‘cascade’ of emails and contacts about the study will be encouraged in each Trust, via staff support teams/leads, chief nursing officers, medical directors, occupational health departments, Union representatives, and wellbeing hub users. The study will be promoted during team briefings, included in Trust newsletters, highlighted by news items on Trust intranet websites, posted to closed social media groups, set by IT services as screen savers on Trust computers, and posters about the study will be displayed in staff rest areas and flyers added in “goodie” bags for staff (in line with any necessary infection control guidelines). Participants will also be recruited via Trust flu clinics.

In Nightingale Hospitals, the study will be promoted, and its importance highlighted, by the staff health and wellbeing team leads at each site, as well as the chief nursing officer, medical director and other team leads involved.

##### Consent

Potential participants will be able to view the information sheet via the study website, then register and provide informed consent if they wish to complete the survey. Paper copies of the participant information sheet (PIS) and consent form can be provided on request. Questions can be emailed to the research team via a dedicated study email address. Participants will provide consent to take part in the study, including baseline and follow up surveys, and can opt-in to consent to be contacted about any linked future studies (e.g. trials of support interventions). We will make clear that participation is voluntary and that participants can withdraw at any time without detriment. Due to the rapid publication of data summaries, it will not be possible to withdraw data from published work, but we will not use withdrawn data in any future publications.

##### Data collection

Baseline data was collected from April 2020 to January 2021. Staff completing the baseline survey will be invited to complete follow up surveys at 6 months and 12 months after they have completed the baseline survey. All participants will be followed up at each subsequent data collection point, regardless of whether they completed the previous survey, unless they choose to withdraw from the study.

The online survey will be administered using Qualtrics survey software, hosted by King’s College London (KCL), and downloaded data will be pseudonymised and stored on secure servers at KCL and University College London (UCL).

#### Diagnostic interview study

##### Recruitment

Participants meeting the inclusion criteria will be emailed with an invitation to participate in the study, including the Participant Information Sheet. Participants who respond to the email indicating their interest will receive a follow up email containing a link to a digital calendar where they can select an interview slot suitable to them using an assigned participant ID, which be confirmed by email by the research team. Participants who do not select a timeslot within 10 days of the booking emails being sent will be prompted to respond if still interested in participating, and followed up by phone calls or texts to confirm interest or withdrawal.

Participants will be able to email the research team via the dedicated study email address with any queries.

##### Consent

1-3 days prior to their interview date, participants will be sent the link to a Qualtrics survey which will contain the online consent form in which they provide the phone number that the research team will call them on for the interview. Provided consent is given, the survey will continue to the administration of the GAD-7, PHQ-9, PCL-6 and GHQ-12.

The morning of the interview, a member of the research team will check completion of the survey; participants who have not started the survey, or who have consented to the study but not completed the measures will be prompted to complete the survey. If necessary, their interview will be rescheduled. Participants who have completed the survey will be send a reminder of their interview time.

##### Data collection

Interviews will be conducted over the phone by researchers based in a confidential location, with calls recorded on an encrypted audio recorder in case of a need for data verification. Researchers will manually enter responses to interview questions into a pre-programmed Qualtrics survey form, following the standard interview procedure for the CIS-R or the CAPS-5 depending on which group the participant belongs to.

Once the interview is complete, the interviewer will email the participant a £25 voucher to thank them for their time and log this payment in the contact database.

#### Qualitative interview study

##### Recruitment

Eligible participants for group i) will be contacted via the email addresses and/or phone numbers provided while completing the baseline and six-month follow up questionnaires. Participants will be invited by email to share their experiences in a qualitative interview. If recruitment is insufficient using email alone participants will be contacted by phone (either phone call or text message) to ascertain interest in the interview study. Recruitment will continue until approximately 12 individuals have been sampled to each category of the sampling frame.

Eligible participants for group ii) will be purposively sampled via existing professional contacts known to the research team. Following initial outreach, snowball sampling will be used until 12 professionals who were involved in the implementation of staff supports in the NHS, have been recruited.

##### Consent

Those expressing an interest in participation will be emailed a PIS and consent form, which they can return via email. Once consent has been obtained, participants will be contacted again to arrange a time suitable to them to be interviewed remotely (via MS Teams/Zoom, or telephone). Interviews are expected to last approximately 30-60 minutes, will be conducted at a time convenient for the participant, and will be recorded with an encrypted Dictaphone. Recordings will later be destroyed following transcription, de-identification, and pseudo-anonymisation of interview transcripts.

##### Data collection

Participants will initially be asked to briefly explain their role and to describe how it did/did not change since the onset of the COVID-19 pandemic. Participants will then be asked about the following: their perceived personal need of support within the NHS and what type of support is needed; their experiences with existing staff support programmes including why they did/did not use them; perceived barriers to access; perceived utility of staff support interventions during the pandemic and in general; other external forms of supports used; suggestions for how supports could be improved in their timing, targeting and content. Before the end of the interview, participants will be given the opportunity to add anything they feel is important that has not been discussed. Participants will receive a £25 Amazon voucher to thank them for their time.

### Study flow chart

Participants will enter the study and be offered the opportunity to participate in different components as outlined in Figure 1.

**Figure 1.**
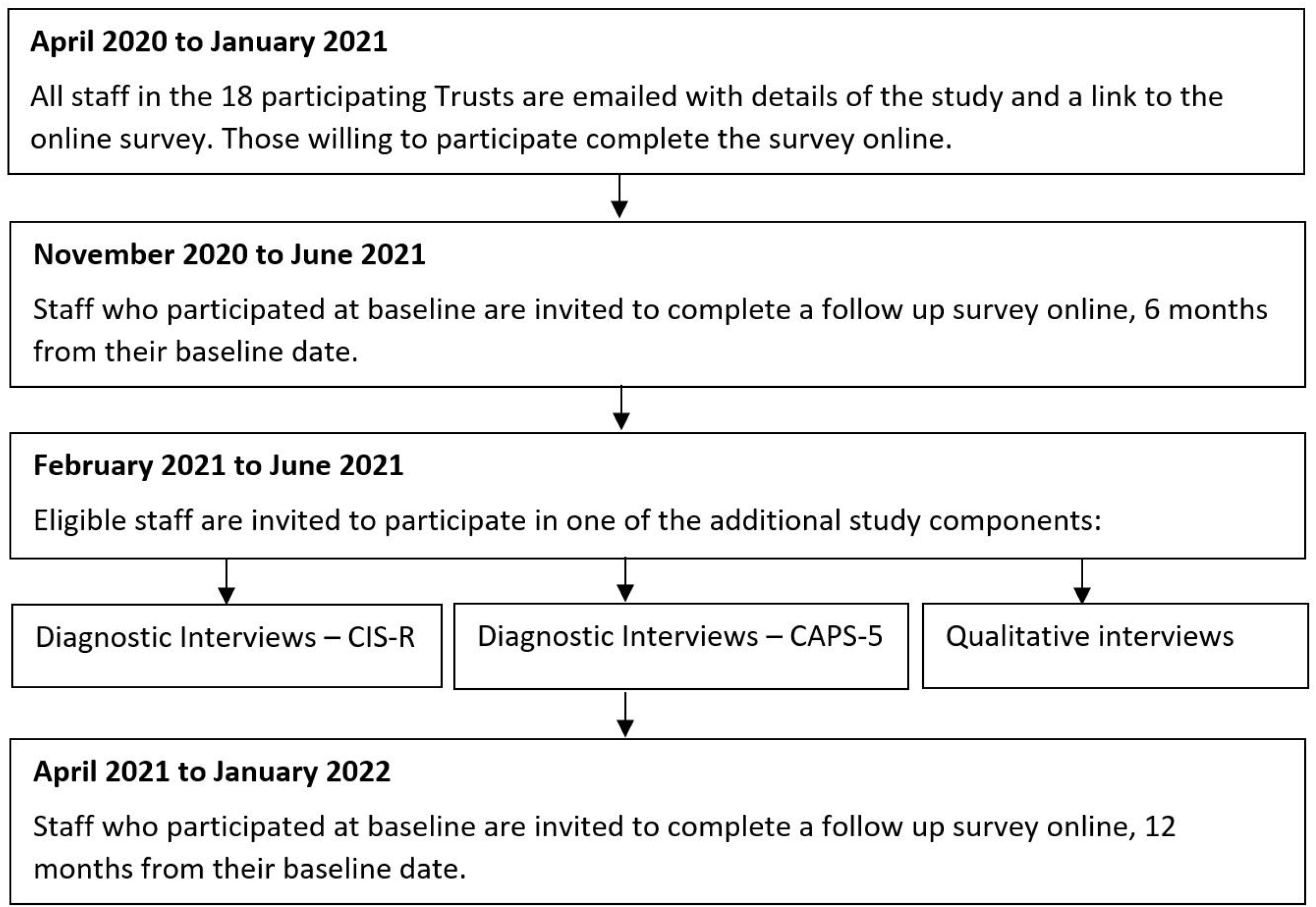
Flow chart of study timings and components

### Analysis plan

#### Longitudinal cohort study

Response weights will be calculated for each Trust, generated using a raking algorithm based on age, sex, ethnicity, and role (using Trust population data obtained from Human Resources), with missing demographic data imputed using multiple imputation (for the purposes of weighting only). Trusts with response rates under 5% will be dropped from the analysis. Representativeness of the sample will be described using frequencies and percentages, and descriptive statistics given for each variable to summarise participants (frequencies and weighted percentages for categorical variables, mean and standard deviation for continuous variables). We will examine differences between participants completing only the short survey, compared to those completing both the short and long surveys. We will summarise the weighted prevalence of the primary and secondary outcomes, stratified by socio-demographic and occupational factors. We will explore potential longitudinal associations between socio-demographic characteristics and occupational factors with the outcome measures (e.g. GHQ-12, GAD-7, PHQ-9, AUDIT, PCL-6, MIES). Three-level random intercept linear or logistic (depending on the distribution of the outcome) regression models will be used to account for the hierarchical structure of the data, considering observations from baseline, 6 month follow up and 12 month follow up at level 2 and NHS trusts at level 3. We will use a measure of pressure on Trusts as the key exposure (e.g. ratio of beds in use to staff numbers, i.e. those not on sick leave), plotting levels of this exposure over time (aggregated at the Trust level), before partitioning the data into meaningful periods corresponding to burden level over the baseline data collection period (April 2020 to January 2021). We will plot all measures over time to look at patterns between this exposure and outcome levels, and use three-level random intercept logistic and linear regression models as appropriate to explore factors associated with outcomes in each period.

#### Diagnostic interview study

This study will validate the screening questionnaires used for general distress and post-traumatic stress disorders using diagnostic interviews with the Clinician Interview Schedule - Revised (CIS-R) and the Clinician Administered PTSD Scale for DSM -5 (CAPS-5) respectively. Participants for the validation studies will be randomly selected from those with non-missing screening scores in a 50% ratio of caseness or otherwise. Sample sizes were obtained by simulation studies (n= 250 and 100 respectively for the CIS-R and CAPS-5 diagnostic interviews). Sensitivity, specificity, positive and negative predictive values will be calculated. If possible, prevalence values for the population at risk will be calculated using population weightings (which will be derived from sex, age and ethnicity variables in an attempt to ensure representativeness in the sample population). Sensitivity analyses will be undertaken to account for missingness if necessary.

#### Qualitative interview study

Interviews will be audio-recorded and transcribed verbatim with identifying information removed. Transcripts will be pseudo-anonymised and uploaded to Nvivo 12 for windows. An inductive qualitative methodology will be used to analyse the interviews underpinned by a pragmatic approach to inquiry. The principles of reflexive thematic analysis will be used to allow an open and organic coding process to develop.[27] Themes will be developed in an iterative process after the initial stages of coding by considering the differences and similarities in the experiences and views of participants from each of the different groups. An initial inductive approach will be applied, more to a more deductive approach over time. A collaborative coding process will be employed, in which members of the research team will initially independently code transcripts and generate a coding framework through discussion. To enhance validity, the emerging thematic framework will be discussed with the wider research team and will be member checked with several interviewees who consented to be contacted again for this purpose at interview.

## Patient and Public Involvement

Frontline NHS staff working in intensive care environments proposed this research, having identified a need to rapidly understand and intervene to try and ameliorate the impact of the pandemic on staff. We tested the proposal’s acceptability and approach with a small informal reference group of front-line staff (psychologists, managers, intensivists and trainee psychiatrists) and refined it accordingly. We have developed an online advisory group of NHS staff (clinical, managerial, auxiliary, students) and NHS patients to provide input on methods development, recruitment strategy, communications, and interpretation of findings. We will also consult this group on tasks such as developing brief lay summaries and interview schedules. We will also run a social media campaign on twitter to raise awareness of the study, as well as to help disseminate our aims, work, and results while restrictions on face to face events are in place. This will be accompanied by a poster campaign to raise awareness, and increase recruitment to the study, with posters being displayed in sites across all the trusts in the study.

## Ethical considerations

### Longitudinal cohort study

Once consented into the study, an automatically assigned ID number will be used for each participant, allowing survey data to be held pseudonymously. All study staff will adhere to relevant data protection regulations, and will maintain confidentiality at all times. No information that could identify any individual participant will be used in reports or publications, or passed on to the Trusts.

Participants will be able to stop the survey at any point, and can skip questions wherever desired. The only required questions are email address and main employing Trust. Signposting information will be provided for any participants who experience distress in answering survey questions (e.g. links to websites for NHS frontline staff, Mind, Samaritans, and the World Health Organisation resources for dealing with psychosocial considerations during the pandemic).

There might be some indirect benefits to the participant when taking part. People often value the opportunity to share their opinions, experiences, and feelings, and several free text options throughout the surveys offer the chance to do this.

### Diagnostic interview study

Participants will be asked to discuss their mental health, which can sometimes cause distress. Participants can stop interviews at any point for a break, postpone the interview to another time or day, or end the interview completely. The researcher will provide immediate emotional support, offer to pause or postpone the interview, and offer to contact a friend, family member, or other supportive person for participants who show signs of distress. The same signposting information outlined above will be provided to any participants who request this. The research team does not take clinical responsibility for research participants in this study; this is made clear during the consent process. However, a standard risk protocol with a supervising clinician will be followed for participants who indicate that they are at risk of harming themselves or others at any stage of the recruitment or participation process.

As with the survey, there may be some benefits to taking part. People often find it helpful talking about their own experiences. The information gained from the study will be used to inform immediate and future responses to the pandemic, and some people enjoy knowing that they have contributed to this.

### Qualitative interview study

Despite the focus of the interview being on evaluating staff supports, some participants may experience distress in answering questions that draw on their experiences of the COVID-19 pandemic. The nature of the interview will be carefully explained in the participant information sheet and the consent form. Participants will be able to pause the interview at any time, skip any questions, and stop the interview entirely, or ask the interviewer to resume another day (this will be arranged wherever possible). Information will be made available at the end of the interview, and after more sensitive questions, for participants who recognise that they are feeling distressed. This will include resources signposting people to support services and helplines. The same risk protocol as above will be followed.

There might be some indirect benefits to the participant when taking part. People often value the opportunity to discuss their experiences and feelings. Further, people may feel keen to contribute to research concerning such a stressful and unprecedented situation. Various members of the research team have been running studies into mental health and wellbeing for many years, including many tens of thousands of participants, and distress resulting from answering questions like the ones proposed in the current study is extremely rare.

## Dissemination

We aim to rapidly disseminate summary findings to the senior management of participating Trusts and collaborating organisations such as NHS England in order to inform staff health and wellbeing strategies. Findings will be more broadly disseminated within the Trusts through their communication channels including websites and staff newsletters. Research findings will also be disseminated to NHS Trusts nationally via our professional network and professional bodies. In addition, findings will be published in academic journals, at conferences and stakeholder meetings and summaries placed on the dedicated study website.

## Data Availability

Access to study data by researchers may be possible on application to the chief investigators.

## Data access

Access to study data by researchers may be possible on application to the chief investigators.

## Contributions

SW, NG, MH, RoR, ReR, and SAMS are chief investigators of the study, contributed to manuscript drafts, and approved the final draft. DL is a co-investigator of the study, and led the write-up of the manuscript. EC, IB and RG wrote the statistical analysis sections of the manuscript, and provided comments on the draft manuscript. RB, HS, SH, and ES are, respectively, the study project manager, post-doctoral researcher, and research assistants, and contributed to the manuscript drafts. AMR, SG, and SM are co-investigators, and provided comments on the draft manuscript. DW is the study data manager, and provided comments on the draft manuscript. RR is the TIDES study link, and provided comments on the draft manuscript.

### The NHS CHECK consortium includes the following co-investigators and collaborators

Sarah Dorrington, Ira Madan, Isabel McMullen, Martin Parsons, Catherine Polling, Danai Serfioti, Chloe Simela, Alexandra Pollitt, Rosie Duncan, Stephani Hatch, Daniel Leightley, Cerisse Gunasinghe, Paul Moran, Peter Aitken, Anthony David, Charlotte Wilson Jones, and Dominic Murphy.

### The NHS CHECK consortium includes the following site leads

Peter Jones, Jeremy Turner, Jesus Perez, Charles Goss, Richard Morriss, Adam Gordon, Frances Farnworth, Julian Walker, Mark Pietroni, Amy Dewar, Sean Cross, Mary Docherty, Scott Weich, David Levy, Ian Smith, Nusrat Husain, Robert Eames, Jessica Harvey, Chris Dickens, Damien Longson, Tayyeb Tahir, Peter Trigwell.

